# The diagnostic certainty levels of junior clinicians: A retrospective cohort study

**DOI:** 10.1101/2020.07.27.20160846

**Authors:** Yang Chen, Myura Nagendran, Yakup Kilic, Dominic Cavlan, Adam Feather, Mark Westwood, Edward Rowland, Charles Gutteridge, Pier D Lambiase

## Abstract

**Purpose of the Study:** To characterise the documentation of working diagnoses and their associated level of certainty by clinicians assessing patients referred to the medical team from the emergency department.

**Design:** This was a single centre retrospective cohort study of non-consultant grade clinicians at the Royal London Hospital, Barts Health NHS Trust between 01/03/19 to 31/03/19. De-identified electronic health record data was collected to include the type of diagnosis documented (clinical, laboratory result or symptom/sign defined) and the certainty adjective used for single diagnoses. Presence or absence of diagnostic uncertainty was collected for multiple diagnoses.

**Results:** 865 medical assessments were recorded during the study period. 850 were available for further analysis. 420 presented a single diagnosis while 430 presented multiple diagnoses. Of the 420 single diagnoses, 67 (16%) were documented as either a symptom or physical sign, and 16 (4%) were laboratory-result defined diagnoses. No uncertainty was expressed in 309 (74%) of the diagnoses. Of 430 multiple diagnoses, uncertainty was expressed in 346 (80%) compared to 84 (20%) in which no uncertainty was expressed.

**Conclusion:** The documentation of working diagnoses is highly variable amongst non-consultant grade clinicians assessing patients admitted via the emergency department. In nearly three quarters of assessments with single diagnoses, no element of uncertainty was implied or quantified. More uncertainty was expressed in multiple diagnoses than single diagnoses however documentation style was heterogenous. These data have implications for prospective studies examining the quantification of diagnostic certainty and its association with important process or outcome measures.

**What is already known on this subject:**

- The factors which influence medical decision-making is almost exclusively assessed by vignettes, simulations or retrospective questionnaires. The certainty or confidence level of a clinician in making a decision can be a source of bias which can lead to patient harm if their confidence is miscalibrated with their accuracy.
- A recent review assessing real world studies of decision-making found only nine, all of which used a Likert or visual analogue scale

**Main messages:**

- The documentation of working diagnoses is highly variable amongst non-consultant grade clinicians
- In nearly three quarters of assessments with single diagnoses, no element of uncertainty was implied or quantified
- Existing documentation is too heterogeneous to meaningfully analyse in a quantitative manner – increased standardisation will allow leveraging of electronic health record platforms to become better educational and research tools for clinicians and educators.

## INTRODUCTION

Clinical medicine is characterised by uncertainty: patients present and manifest pathology in a myriad of ways. Most clinical research focuses on creating an evidence base to support the safe and effective use of new or existing diagnostics and treatments. However, the decision making processes required to select the optimal clinical strategy has not been assessed as rigorously. Accurate decision-making represents a key step in providing high quality healthcare to patients.^1^

One important factor that may influence the decision to select a particular investigation or treatment is the degree of certainty a clinician feels regarding his or her working diagnosis. Such certainty may be affected by a multitude of both internal and external factors as well as a lack of complete data when making decisions.^2^

The act of diagnostic calibration is the process by which a clinicians’ confidence in the accuracy of their diagnosis aligns with their actual accuracy.^3-5^ This alignment of confidence and accuracy requires that both are precisely measured. For the former, the explicit measurement of confidence or certainty has been predominantly assessed in controlled environments, such as retrospective questionnaires after the clinical interaction, vignette studies or in simulations.^6-9^

Better alignment of confidence and accuracy could mitigate against errors related to hubris or devaluing of underconfident opinions^10^, and in a 2015 Institute of Medicine report it was noted that “nearly all patients will experience a diagnostic error in their lifetime, sometimes with devastating consequences”.^11^

A recent systematic review focusing solely on decision certainty measured in real-time captured only nine studies – all of which used a scale such as a Likert or visual analogue scale.^12^

Our study therefore had two main research aims

1. To characterise the documentation of diagnoses by clinicians in a real world setting of an acute medical admission (the medical take)
2. To highlight the distribution of self-rated certainty amongst documented diagnoses and to suggest relevant follow-on research questions to test hypotheses generated as a result of this study.

## METHODS

The study protocol was registered with the Clinical Effectiveness Unit at Barts Health NHS Trust (ID no. 10201). Permission from the Chief Clinical Information Officer was granted to conduct a service evaluation project assessing the documentation of patients admitted through the medical take at the Royal London Hospital by clinicians using an electronic health record (EHR), called Cerner Millennium. All data were stored on a secure server used by Barts Health NHS Trust for quality improvement and research studies. The manuscript has been prepared according to The Strengthening the Reporting of Observational Studies in Epidemiology (STROBE) Statement.^13^

### Data sources

A standardised proforma was used by all clinicians when assessing patients referred to the medical team (clerking proforma). During the data collection period, this was mandated as part of local departmental governance at the Royal London Hospital (RLH). The specific field of interest in our study was the ‘working diagnosis’ (see figure 1). EHR data at the Royal London Hospital can be accessed by Cerner Business Objects – a service evaluation tool^14^ used by Barts Health NHS Trust – which provides a graphical user interface to analyse data within the EHR and run SQL queries. We extracted all clerking proformas recorded in March 2019 and cross-checked the list against a manual database of admissions maintained by the acute medicine department under the medical take. Extracted clerkings were anonymised and then exported to Microsoft Excel where a regular expression function was used to crop the working diagnosis from each clerking.

**Figure 1.**
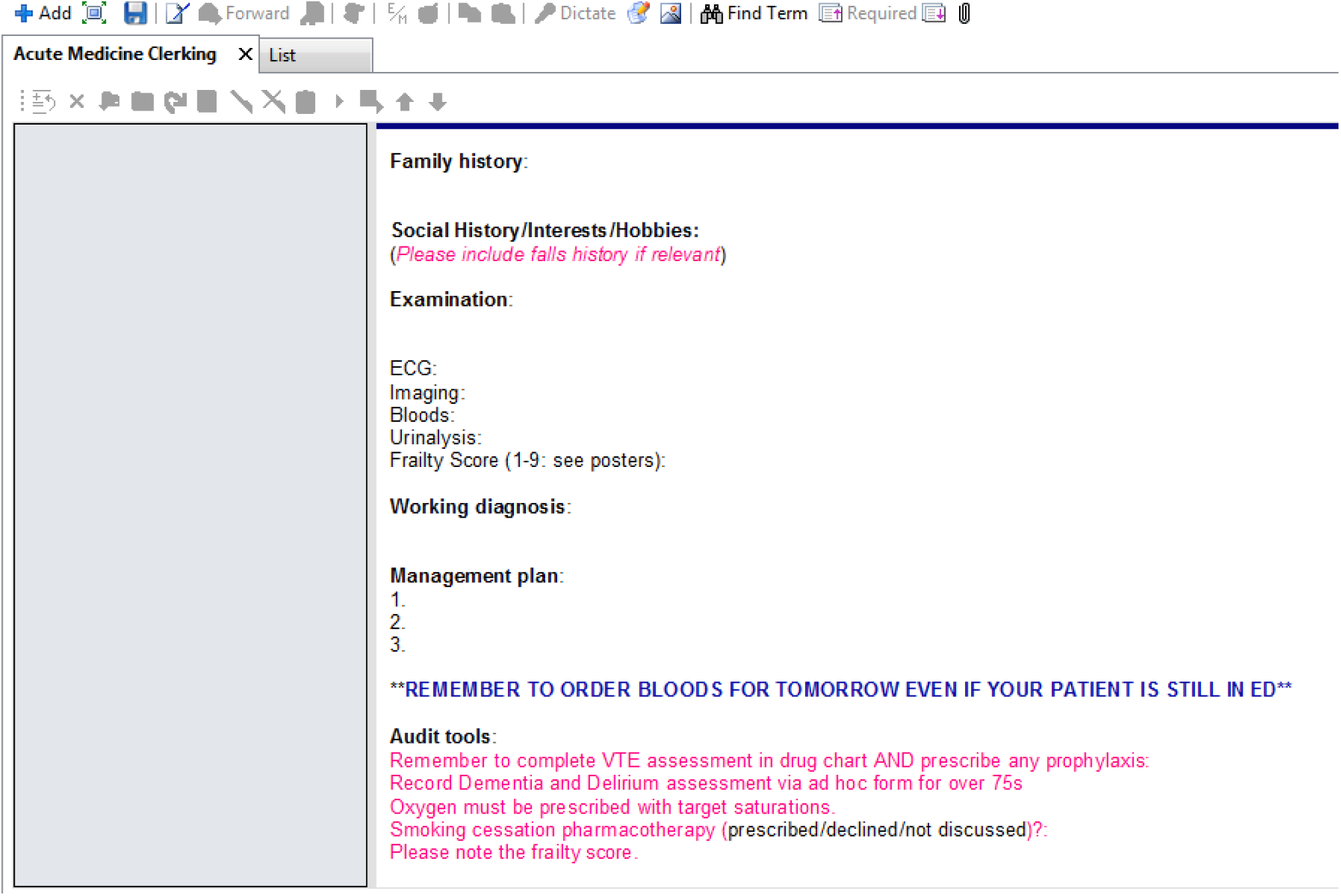
Appearance of clerking proforma to clinicians at study site

### Hierarchy of certainty, definition of diagnosis and diagnosis type

Two clinicians each with 7 years post-graduate clinical experience (YC & MN) along with expert input from three clinicians with >50 years combined post-graduate clinical experience (DC, AF & PL) agreed upon the classification of terms documented in the ‘working diagnosis’ field, grouped by the degree of uncertainty associated (see Figure 2).

**Figure 2.**
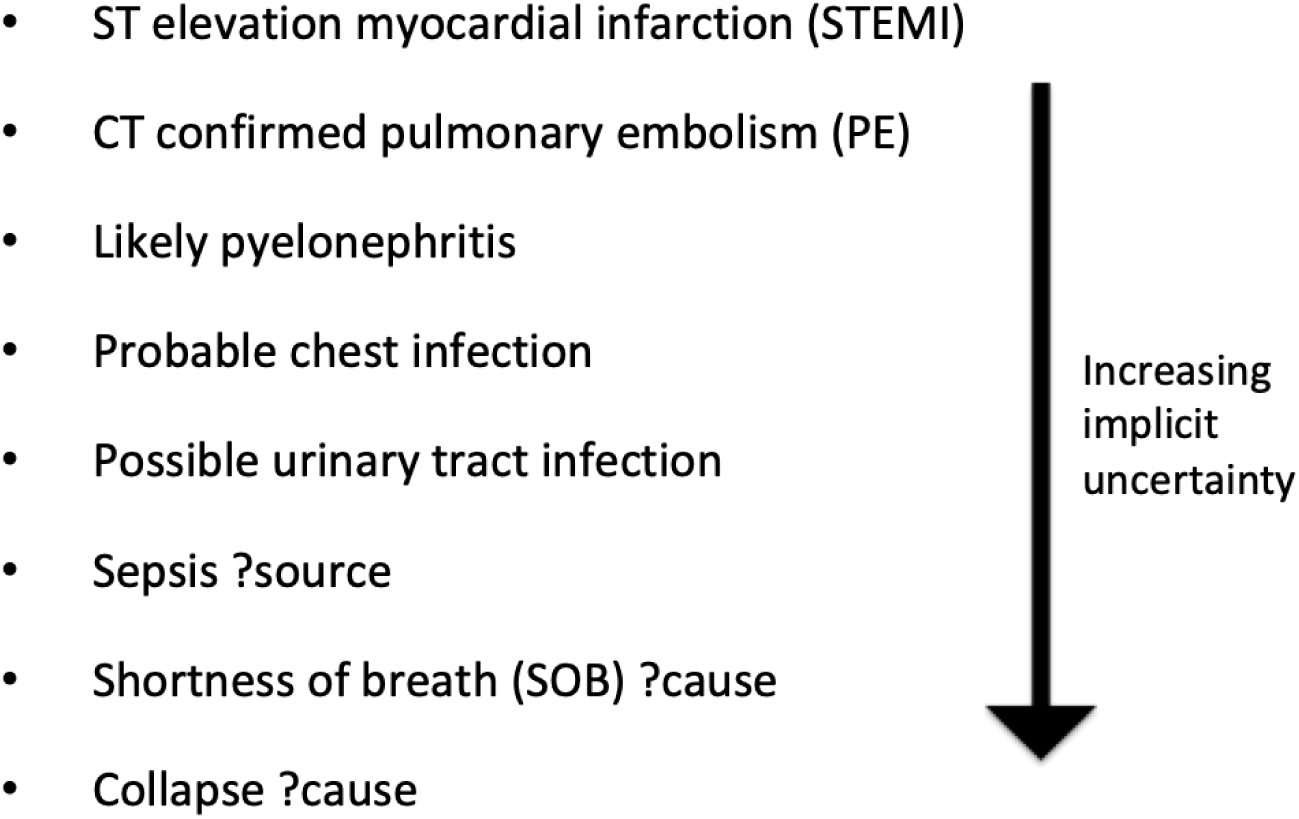
Examples of terms used in study sample and associated ranking of certainty.

A diagnosis was defined as whatever the clinician had documented at the time of assessment in the ‘working diagnosis’ section, and was subject to patient factors (complexity of the case) as well as individual factors (documentation style). Many were strictly speaking, not clinical diagnoses in the traditional medical sense; some were even repetitions of the presenting complaint.

All ‘working diagnoses’ were split into two initial categories: single diagnoses and multiple diagnoses. For single diagnoses, categorisation of certainty was made on the basis of the specific verbal adjectives or descriptors present. This categorisation was modelled on high stakes decision-making in the face of uncertainty in other settings (see Table 1).^15^

**Table 1.** Categorisation of certainty, based on National Intelligence Community in the US.

|  | No<br>qualifier | Definitely<br>not | Very unlikely | Less likely | Likely | Very likely | Definite |
| --- | --- | --- | --- | --- | --- | --- | --- |
| Synonyms | n/a | Excluded | Doesn't, rule<br>out, need to<br>exclude | ?, Query,<br>Possible,<br>Consider | Probable | More likely | Confirmed |
| %<br>likelihood<br>of stated<br>condition | n/a | 0 | 0-20 | 20-55 | 55-80 | 80-99 | 100 |
| Examples | Septic<br>shock<br>secondary<br>to CAP<br>NSTEMI | HIV<br>excluded<br>by<br>negative<br>serology<br>Mumps<br>excluded<br>as has<br>been vaccinated | Inflammatory<br>disease,<br>aetiology<br>unclear. Rule<br>out TB<br>Need to<br>exclude VP<br>shunt<br>infection | ?viral illness<br>Consider<br>paracetamol<br>overdose<br>Possible<br>subarachnoid<br>haemorrhage | Likely<br>gastritis<br>causing<br>vomiting<br>Probable<br>sepsis | Acute<br>confusion,<br>very likely<br>to be either<br>overdose of<br>prescription<br>medication<br>or<br>recreational<br>drug use | Confirmed<br>PE on<br>CTPA<br>Definite<br>STEMI |

Single diagnoses were also subdivided based on the hierarchy of diagnosis level. For example, the lowest level was a symptom or sign based diagnosis e.g. chest pain. In cases where the documented diagnosis was relaying a patients’ reported chest pain, the expectation was that this would carry no certainty descriptor or for certainty to be definite. The level above this was a diagnosis based on a laboratory result e.g. hyperkalaemia. In such cases, the expectation was once again that this would carry no certainty descriptor or for certainty to be definite. These two levels were contrasted against clinical diagnoses of conditions e.g. unstable angina, where the expectation was that a certainty qualifier would be attached.

For multiple diagnoses, a more basic analysis was conducted, after discussion between the authorship group revealed differences in the interpretation of the written record and difficulties in allocating such records to certainty descriptors for each multiple diagnosis. It was recognised that this group of results were at particular high risk of bias.

Table 2 outlines examples of these complex records:

**Table 2.** Examples of complexity of documentation of multiple diagnoses on the clerking proforma

| **Sample working diagnosis from assessments** | **Are diagnoses separate or are some layered / differentials/ contain triggers or sequelae?** | **Was uncertainty expressed?** |
| --- | --- | --- |
| Reduced GCS 9/15 - ?cause, ?medication-related - coincided with clozapine<br>Hypertension - BP ~200 systolic<br>Less likely neuroleptic malignant syndrome - rare on clozapine, high CKs but no fever/rigidity/raised WCC | Some separate diagnoses, some are potential differentials | Yes |
| Fall - multifactorial poorly controlled PD<br>deterioration in patient's mobility catheter associated UTI | Some separate | No |
| LRTI on B/G of COPD<br>Multifactorial fall due to postural hypotension (secondary to drugs and dehydration), frailty --> need to rule out silent cardiac event as patient has had a history of this.<br>--> not in keeping with AS being cause of syncope<br>AKI on CKD likely due to poor oral intake | Some separate | Yes |
| HAP<br>Frailty<br>Poor oral intake - dry | Separate, but some descriptors may relate to co-morbidities and background | No |
| Difficult diagnosis<br>Most likely sepsis ?source with high lactate<br>Chest, abdominal, CNS<br>Acute behavioural disturbance, on antipsychotics, no obvious history of substance abuse<br>NMS - essentially normal peripheral neurological examination - unlikely | Some separate, some are potential differentials | Yes |

Thus multiple diagnoses either represented more than one condition being manifest in that particular clinical case or represented the explicit recognition of diagnostic uncertainty through differential diagnoses. Additionally, some records pertained to layering of diagnoses – for instance secondary diagnoses that were sequelae or triggers of the main diagnosis. These also proved difficult to analyse in a systematic manner. Therefore we categorised multiple diagnoses as either containing any expression of uncertainty within them or not.

### Patient and public involvement (PPI)

A focus day was held in March 2019 to inform the study question and design, assessing the views of patients regarding clinicians’ diagnostic certainty level. 11 patients (7 female, 4 male) attended the session lasting approximately one and a quarter hours. Both before and after a discussion session, patients were asked “how important to patients and the public do you think it is to conduct this research?”. The response scale ranged from 1 (not at all important) to 5 (very important). A mean score of 4.5 increased to 4.9 after an open forum discussion session. There was unanimous support for more research to be conducted on the topic of uncertainty in working diagnoses for medical patients.

## RESULTS

865 clerking proformas were analysed. Automated data extraction was 100% complete when cross-referencing against the manual database. 15 records were unsuitable for further analysis (9 had no diagnosis recorded, 3 could not be extracted from the electronic health record due to technical issues and 3 were elective as opposed to acute admissions). This left 850 available for further analysis of which 420 presented a single diagnosis while 430 presented multiple diagnoses (Figure 3).

**Figure 3.**
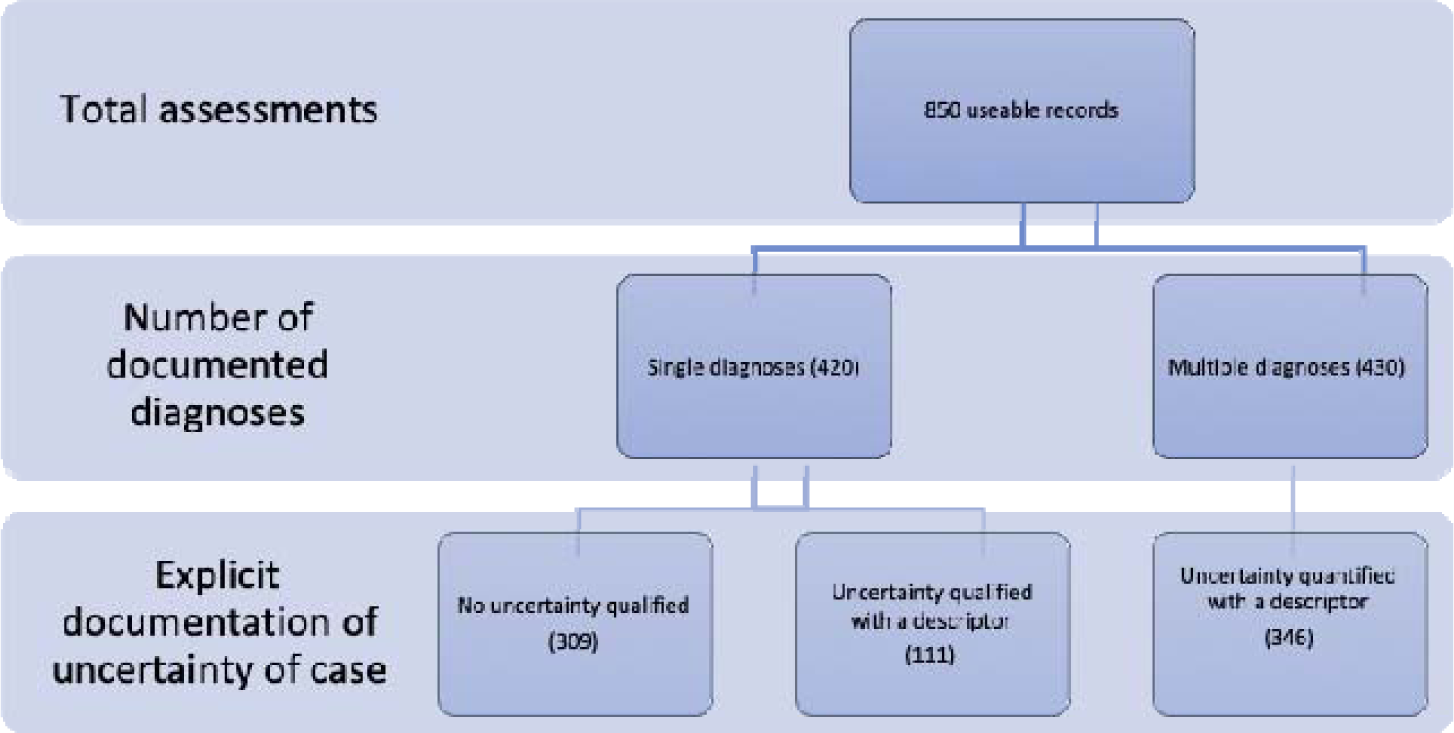
Summary of documentation of assessment, categorised by number of diagnoses

Of the 420 single diagnoses, 67 (16%) were symptom or sign defined while 16 (4%) were laboratory result defined diagnoses. The remaining 337 (80%) were diagnoses of a specific condition. There was no verbal adjective or quantification of uncertainty in 74% of the single diagnoses (309 of 420) and no diagnosis was described as definite, certain, confirmed or excluded. The description of certainty in the remaining 111 cases was categorised according to the classification system in Table 1 as follows: very unlikely (16), less likely (58), likely (31), more likely (6).

Of the 430 multiple diagnoses, uncertainty was expressed in 346 (80%) compared to 84 (20%) in which no uncertainty in the diagnoses was expressed. Examples of multiple diagnoses are outlined in Table 2.

In total, 71 different clinicians contributed to the assessment of medical patients during the study period. Their names and grades were not available for analysis as part of this service evaluation, however, the clinical staffing model at RLH would allow for an approximation of a 4:1 ratio of ‘Senior House Officer’ grade clinicians (between 1-4 years of postgraduate experience) to Registrar grade clinicians (5+ years of postgraduate experience) in this sample.

## DISCUSSION

To our knowledge, this study is the first real world analysis of how uncertainty in the working diagnosis is documented at an acute medical hospital in the UK. There are four key findings. Firstly, approximately half of working diagnoses contain only a single diagnosis as opposed to a differential. Secondly, in nearly three quarters of single diagnoses, no element of uncertainty is documented – this may relate to the way UK clinicians are trained or may reflect cases that are straightforward. Thirdly, a fifth of single diagnoses are defined entirely by a symptom/physical sign or laboratory result rather than generation of a clinical diagnosis. Lastly, characterising multiple diagnoses into more granular categories was predictably difficult, owing to the inherently varied ways in which they are recorded. The recording of uncertainty amongst non-consultant grade clinicians appears ad-hoc, implicit and discretionary, making quantitative analysis and designing future quality improvement efforts difficult.

### Comparison with the literature

Decision making research has focused on surveys undertaken away from the frontline or simulated scenarios using case vignettes – this allows for careful control of multiple variables that may influence the results. The paucity of research examining decision making in real world settings has been because of a perceived lack of ability to control for obvious variables, and to be able to meaningfully link a snapshot decision to any hard outcomes such as length of stay or inpatient mortality.

However, our contention is that there is value in pursuing this line of research even if it is difficult. There are a number of beneficial second-order effects that could arise through more robust and meaningful pursuit of analysing real-world decision making. For example, there may be educational value, both for trainees and senior doctors – and the wider workforce – in seeing how their self-rated certainty relates to longer-term outcomes, along with how they compare to their peers. There is evidence that already exists for the benefits of an embedded audit and feedback system that facilitates reflective practice for staff.^16^

In a detailed mixed methods study by O’ Hara et al,^17^ staff raised concerns regarding the impact of external pressures on decision-making such as admission avoidance guidance: “You are almost going into a ridiculous level of risk management, which actually isn’t to do with patient care.” The study of decision-making and how it relates to internal and external pressures will rely upon more systematic measurement of decision-making certainty which is currently open to interpretation – the meaning of what ‘likely’ or ‘probable’ is will be different to different people and has been the subject of decades of behavioural science, sociology and psychology research.^18^

### Follow up

There are three parallel questions which emerge from this study:

1. Can we prospectively measure certainty levels in a more robust manner and are they associated with process and outcome measures?
2. To what extent is certainty attributable to the patient versus external and internal factors; e.g. simple vs. complex patient, a night vs. day shift, a clinicians’ seniority or even their personality/ style of thought processes?
3. Can we design cognitive interventions to mitigate against mis-calibrations between diagnostic accuracy and diagnostic confidence?

One potential next step would be to introduce a Likert scale within the EHR asking the clinician to rate their certainty level for the working diagnosis. Quantification would allow easier testing of pragmatic research questions. Correlating a diagnostic forecast to its accuracy using traditional measures such as an area under the curve (AUC) of a receiver operating characteristic or a Brier score will require carefully designed studies to avoid biases such as misplaced conclusion of poor accuracy when diagnoses overlap or outcomes and management strategies relate to one another.^19^

Of note, this is a subjective area. One doctors’ perception of 90% certainty will be different to that of another. But this in itself contains valuable information and insights from an educational perspective. Appreciating the variation amongst clinicians’ thought processes represents an important antithesis to better understand, when compared to the standardisation and ‘removal of unwarranted variation’ advocated by guidelines, standard operating procedures, and increasingly, artificial intelligence algorithms.

In any future work, care must be taken not to add any further burden onto the already stretched clinical workflow. Bassford et al^20^ demonstrated that adding a standardised referral form when deciding to admit a patient to intensive care had barriers to uptake. However, of the one third of clinicians who made use of the form, there was evidence supporting an impact on decision-making: clinicians noted that the forms had prompted them to consider blind spots in their thinking including a greater focus on the views of the patient.

### Implications

Explicit self-rating of certainty could directly contribute to improving patient safety and advocacy through the Hawthorne effect. The mere act of quantifying a certainty rating might form a cognitive brake and thus an important debiasing strategy that mitigates against avoidable medical error occurring from under- or over-confidence.^21-25^ Patients in our PPI work agreed that low certainty ratings could help identify difficult cases and prompt greater teamwork. It is conceivable that improvements in clinical processes such as reduced waste from over-investigation or better patient flow to appropriate environments could arise as a result of highlighting patients where decisions are made with high or low certainty. In a systematic review of real-time decision making, one study looked for a correlation between certainty and clinical outcomes: a high uncertainty for the diagnosis of heart failure was associated with a longer length of stay, increased mortality and higher readmission rates at 1 year.^12^ Future research will likely examine prospective testing of artificial intelligence (AI) decision support tools in healthcare and a logical question to test is how clinicians will interact and engage with such tools i.e. what factors (including certainty levels) influence whether clinicians agree or disagree with AI-recommended management decisions.

### Limitations

Our findings must also be considered in light of several limitations. First, due to logistical constraints, we were unable to link the working diagnosis to the final discharge diagnosis to assess how documented uncertainty relates to diagnostic accuracy. Nor were we able to link uncertainty to process measures or outcomes. Any analysis which attempts to link snapshot diagnostic decision-making at the start of a patient’s disease course, with the ultimate diagnosis at discharge, will be heavily caveated by the passage of time. In one UK study^26^ only 12% of diagnoses made by referring clinician matched the final diagnosis made after subsequent work up in hospital.

Second, we were not able to analyse the multiple diagnoses with the same granularity as the single diagnoses. We therefore made the pragmatic decision to approximate uncertainty by tagging the record as uncertain if there were any of the descriptors from Table 1 present. This may explain why a larger proportion of multiple diagnoses contained some degree of uncertainty.

Third, owing to the modest sample size, we did not explore other relevant external factors which may have influenced decision certainty, including the time of day and seniority of clinician – the latter being a factor identified in a vignette study^8^ as a surrogate for greater tolerance of uncertainty in an emergency department setting. Additional factors such as the complexity of the patient and their acuity of presentation, as well as the comprehensiveness of the initial ED referral and assessment would likely have significant effects on the documentation of certainty of working diagnosis.

Finally, the sample arises from a single teaching hospital and may not be representative of other institutions, particularly in international settings where educational or cultural differences may have a significant impact on clinician self-rating of certainty.

## Conclusions

In nearly three quarters of single diagnoses, no element of uncertainty is portrayed or quantified. Greater uncertainty is expressed in multiple diagnoses than single diagnoses. These data have implications for the design of prospective studies looking to assess how uncertainty is recorded and whether there is an association between certainty of working diagnosis and process measures or outcomes.

## Data Availability

Raw anonymised data is available from the corresponding author upon reasonable request.

## Author Contributions

YC and MN conceived the idea. YC and MN extracted the data. YC, MN and YK curated and analysed the data. DC designed the clerking proforma. DC, AF and PL arbitrated on the hierarchy of certainty. MW, ER and CG contributed and edited the original service evaluation proposal. YC wrote the first draft of the manuscript and all authors contributed to subsequent revisions. All authors approved the final manuscript. YC is the guarantor and takes responsibility for the integrity of the work and confirms that the decision on behalf of the authors to publish this work.

